# Risk Factor Analysis for Cancer and Coronary Heart Disease: A Machine Learning Approach Using National Health and Nutrition Examination Survey Data

**DOI:** 10.1101/2024.11.05.24316754

**Authors:** Bozcuk Hakan Şat

## Abstract

**Objectives:** The relative significance of predictive factors for cancer and coronary heart disease (CHD) is still unclear. This study aims to identify and evaluate the risk factors contributing to the development of both conditions using the CatBoost machine learning algorithm.

**Methods:** Data from twelve datasets of the 2009–2010 National Health and Nutrition Examination Survey (NHANES), incorporating both survey responses and laboratory results, were used. Separate CatBoost models were developed to predict cancer and CHD occurrences, by using Shapley Additive Explanations (SHAP), with the help of Recursive Feature Elimination with Cross-Validation (RFECV), and by adjusting class weights, and model performance was assessed using Receiver Operating Characteristic (ROC) curves.

**Results:** The datasets were combined to form a cohort of 5,012 participants, each with 24 selected features. The cancer prediction model achieved a ROC Area Under the Curve (AUC) of 0.76, with 13 selected features, yielding an accuracy of 0.70, sensitivity of 0.67, and specificity of 0.70. In contrast, the CHD prediction model achieved a higher ROC AUC of 0.87, with an accuracy of 0.83, sensitivity of 0.78, and specificity of 0.83. Accordingly, top predictive features for each disease have been ranked and selected by the CatBoost algorithm.

**Conclusions:** This study identifies key demographic and laboratory features significantly associated with cancer and CHD risk in the NHANES dataset. The findings suggest that these factors could be valuable for estimating individual risk and could inform machine learning models aimed at early detection and screening.

## INTRODUCTION

Cancer and coronary heart disease (CHD) are two major causes of morbidity and mortality worldwide, and they share several common etiological factors. Both conditions are particularly prevalent in older populations [1–3]. While numerous predictive factors have been identified for each disease individually, the exact mechanisms through which these factors interact, and the specific pathways involved in the simultaneous development of both cancer and CHD, remain incompletely understood. There is a subset of patients who develop both diseases concurrently, yet the specific predisposing factors that drive the co-occurrence of these conditions remain elusive, leaving a significant gap in our knowledge of the underlying mechanisms and risk factors.

Machine learning is increasingly being applied across various medical disciplines and has shown great promise in transforming clinical care. By using tabular, image, and sound data, machine learning algorithms are capable of detecting patterns that can aid in diagnosing diseases, predicting prognoses, and selecting treatment plans [4,5]. These algorithms have already demonstrated substantial utility in applications such as breast cancer diagnosis, personalized treatment plans based on genetic profiles, and correlating brain function with imaging findings from Positron Emission Tomography/Computed Tomography (PET/CT) in non-small cell lung cancer [6–8]. Moreover, machine learning has been successfully employed to predict short-term outcomes in spontaneous intracerebral hemorrhage by identifying key predictors, demonstrating its valuable potential in prognostic models [9].

Among the various machine learning algorithms, CatBoost (short for Categorical Boosting) stands out as a particularly powerful gradient boosting algorithm. Developed by Yandex, CatBoost is user-friendly and often requires less parameter tuning than other gradient boosting algorithms, such as XGBoost or LightGBM [10].

Within the National Health and Nutrition Examination Survey (NHANES) dataset, certain subgroups have been diagnosed with either cancer, CHD, or both [11]. However, it remains unclear whether the factors that predict the development of cancer and CHD overlap or are distinct. Therefore, the aim of this study is to identify the predictors of cancer and CHD using comprehensive data from the NHANES survey and to evaluate the potential of the CatBoost machine learning algorithm in this task. By leveraging the NHANES dataset and applying CatBoost, I seek to uncover key predictive features for each disease, aiming to support early detection and intervention strategies.

## METHODS

### General Issues, Collection, and Processing of Data

The open-access database from the National Health and Nutrition Examination Survey (NHANES) was obtained from its official website at the following web address: “https://www.cdc.gov/nchs/nhanes/index.htm” [11]. For this study, focusing on the period of 2009– 2010, twelve datasets were downloaded from the NHANES website. These datasets included ALQ_F.XPT (for alcohol use), CBC_F.XPT (for complete blood count), WHQ_F.XPT (for weight history), SMQ_F.XPT (for smoking and cigarette use), PAQ_F.XPT (for physical activity), MCQ_F.XPT (for medical conditions), INQ_F.XPT (for income), CRP_F.XPT (for C-Reactive Protein; CRP), GHB_F.XPT (for Glycosylated Hemoglobin; HbA1c), DBQ_F.XPT (for diet and nutrition), TCHOL_F.XPT (for cholesterol), and DEMO_F.XPT (for demographics). Initially, these datasets were in XPT format, which were subsequently converted into Pandas data frames for further analysis.

The next step involved filtering each dataset to include only the common SEQN values across all databases. SEQN, or “Sequence Number,” serves as a unique identifier assigned to each participant in the survey. By focusing on the SEQN values present in all datasets, a combined dataset was generated that contained features from each database. This combined dataset was further processed by removing rows with any missing values, ensuring that only complete cases were included for the analysis. This clean, processed dataset was then saved in Excel format for subsequent steps. All data processing was conducted in Google Colab, utilizing Python programming, with the Pandas and Openpyxl libraries [12–14].

The variables tested as potential predictors in this study encompassed a wide range of demographic, behavioral, and biological factors. These included age, gender, alcohol consumption, smoking history, dietary habits, body weight, physical activity level, income, family history of heart attack, previous cancer diagnosis, CHD diagnosis, white blood cell count, percentages of lymphocytes and neutrophils, hemoglobin levels, red cell distribution width, platelet count, mean platelet volume, CRP, total cholesterol, and HbA1c. These variables were selected due to their relevance to the development of cancer and CHD, their possible links to chronic inflammation, and accessibility in the NHANES database.

This draft of the study was initially linguistically enhanced with the assistance of ChatGPT 4.0, but the final version of the article was carefully reviewed and approved by the author.

### Development of CatBoost Models to Predict Cancer or CHD

The descriptive analysis for this study was conducted using SPSS version 21.0.0 [15]. In the second phase, the CatBoost machine learning algorithm was employed to develop predictive models for the occurrence of cancer and CHD. Considering that the cancer and CHD subgroups comprised only a minority of the total cohort, I specifically adjusted the class weights in the CatBoost model to account for the imbalance between the minority and majority subgroups. This adjustment aimed to ensure that the model could learn effectively from the minority class data.

To select the most important predictive features, I made use of the Recursive Feature Elimination with Cross-Validation (RFECV) algorithm [16]. The RFECV algorithm is a feature selection method that iteratively removes the least important features from a model to find the most relevant subset of predictors. It starts by training a model using all features and ranks their importance based on a specified metric, such as feature weights or coefficients. The least important feature is then removed, and the model is retrained. This process repeats until a predefined number of features remain or performance plateaus. Cross-validation is used to ensure the selected features improve the model’s performance in a robust manner, preventing overfitting and making the selection process more reliable. In this study, in order to optimize the set of predictive features, I manually adjusted the parameters of the RFECV algorithm; particularly “select” and “min_features_to_select”, using F1 score for scoring.

The F1 score, as I used for scoring the RFECV algorithm in this study, is a performance metric used in classification that balances precision and recall [17]. It is the harmonic mean of these two metrics, making it particularly useful when the classes are imbalanced. It is calculated as:

F1 = 2 x ((Precision × Recall) / (Precision + Recall)).

Feature importance rankings were calculated based on the dataset, and Shapley Additive Explanations (SHAP) values were computed and visualized to evaluate the contribution of each feature to the predictions [18]. Additionally, confusion matrices were generated to assess model performance, along with key evaluation metrics, such as accuracy, sensitivity, and specificity. Receiver Operating Characteristic (ROC) curves were also plotted, and the Area Under the Curve (AUC) values were calculated to assess the overall predictive performance of the models for both cancer and CHD [19].

This second CatBoost analysis phase of the study was also completed using Google Colab, with the analysis performed in Python, utilizing several libraries, including Numpy, Pandas, Scikit-learn, CatBoost, and Matplotlib [12,20–23]. These tools allowed for the efficient handling of the dataset and the development of accurate machine learning models to predict cancer and CHD risk based on the selected features.

## RESULTS

### General Findings

A total of 5,012 cases were analyzed, with a gender distribution of 50.3% female and 49.7% male. Additionally, 55.3% of the cases were categorized as overweight. Of the total sample, 4,343 cases (86.7%) had no diagnosis of either cancer or CHD, while 462 cases (9.2%) were diagnosed with cancer, and 157 cases (3.1%) were diagnosed with CHD. Co-occurrence of both cancer and CHD was observed in 50 cases (1%). Laboratory values for the participants were generally within normal ranges, with some variability. For example, the mean hemoglobin level was 14.1 g/dL (standard error = 1.5). The demographic characteristics, along with other features derived from questionnaires and laboratory tests, are presented in Table 1.

**Table 1.**
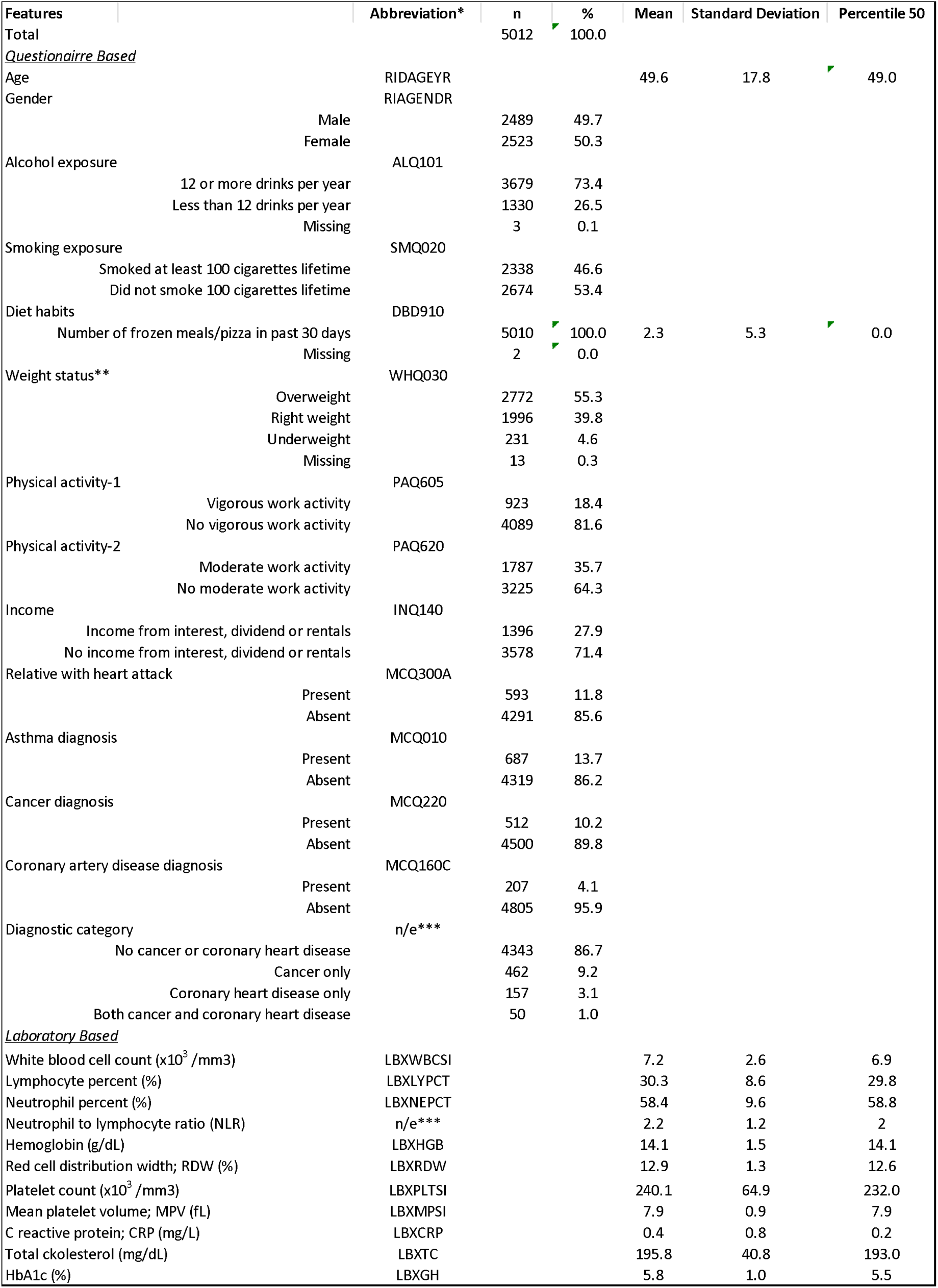
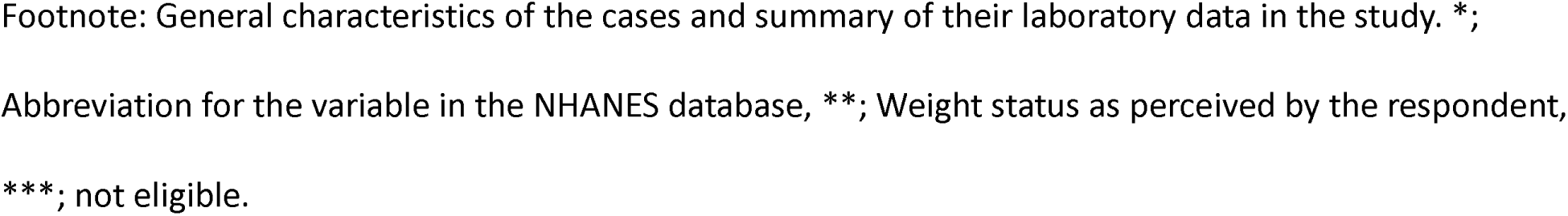
Demographics, questionnaire and laboratory-based data from NHANES study (2009-2010).

| Features | Abbreviation* | n | % | Mean | Standard Deviation | Percentile 50 |
| --- | --- | --- | --- | --- | --- | --- |
| Total |  | 5012 | 100.0 |  |  |  |
| *Questionnaire Based* |  |  |  |  |  |  |
| Age | RIDAGEYR |  |  | 49.6 | 17.8 | 49.0 |
| Gender | RIAGENDR |  |  |  |  |  |
| Male |  | 2489 | 49.7 |  |  |  |
| Female |  | 2523 | 50.3 |  |  |  |
| Alcohol exposure | ALQ101 |  |  |  |  |  |
| 12 or more drinks per year |  | 3679 | 73.4 |  |  |  |
| Less than 12 drinks per year |  | 1330 | 26.5 |  |  |  |
| Missing |  | 3 | 0.1 |  |  |  |
| Smoking exposure | SMQ020 |  |  |  |  |  |
| Smoked at least 100 cigarettes lifetime |  | 2338 | 46.6 |  |  |  |
| Did not smoke 100 cigarettes lifetime |  | 2674 | 53.4 |  |  |  |
| Diet habits | DBD910 |  |  |  |  |  |
| Number of frozen meals/pizza in past 30 days |  | 5010 | 100.0 | 2.3 | 5.3 | 0.0 |
| Missing |  | 2 | 0.0 |  |  |  |
| Weight status** | WHQ030 |  |  |  |  |  |
| Overweight |  | 2772 | 55.3 |  |  |  |
| Right weight |  | 1996 | 39.8 |  |  |  |
| Underweight |  | 231 | 4.6 |  |  |  |
| Missing |  | 13 | 0.3 |  |  |  |
| Physical activity-1 | PAQ605 |  |  |  |  |  |
| Vigorous work activity |  | 923 | 18.4 |  |  |  |
| No vigorous work activity |  | 4089 | 81.6 |  |  |  |
| Physical activity-2 | PAQ620 |  |  |  |  |  |
| Moderate work activity |  | 1787 | 35.7 |  |  |  |
| No moderate work activity |  | 3225 | 64.3 |  |  |  |
| Income | INQ140 |  |  |  |  |  |
| Income from interest, dividend or rentals |  | 1396 | 27.9 |  |  |  |
| No income from interest, dividend or rentals |  | 3578 | 71.4 |  |  |  |
| Relative with heart attack | MCQ300A |  |  |  |  |  |
| Present |  | 593 | 11.8 |  |  |  |
| Absent |  | 4291 | 85.6 |  |  |  |
| Asthma diagnosis | MCQ010 |  |  |  |  |  |
| Present |  | 687 | 13.7 |  |  |  |
| Absent |  | 4319 | 86.2 |  |  |  |
| Cancer diagnosis | MCQ220 |  |  |  |  |  |
| Present |  | 512 | 10.2 |  |  |  |
| Absent |  | 4500 | 89.8 |  |  |  |
| Coronary artery disease diagnosis | MCQ160C |  |  |  |  |  |
| Present |  | 207 | 4.1 |  |  |  |
| Absent |  | 4805 | 95.9 |  |  |  |
| Diagnostic category | n/e*** |  |  |  |  |  |
| No cancer or coronary heart disease |  | 4343 | 86.7 |  |  |  |
| Cancer only |  | 462 | 9.2 |  |  |  |
| Coronary heart disease only |  | 157 | 3.1 |  |  |  |
| Both cancer and coronary heart disease |  | 50 | 1.0 |  |  |  |
| *Laboratory Based* |  |  |  |  |  |  |
| White blood cell count (x10<sup>3</sup> /mm3) | LBXWBCSI |  |  | 7.2 | 2.6 | 6.9 |
| Lymphocyte percent (%) | LBXLYPCT |  |  | 30.3 | 8.6 | 29.8 |
| Neutrophil percent (%) | LBXNEPCT |  |  | 58.4 | 9.6 | 58.8 |
| Neutrophil to lymphocyte ratio (NLR) | n/e*** |  |  | 2.2 | 1.2 | 2 |
| Hemoglobin (g/dL) | LBXHGB |  |  | 14.1 | 1.5 | 14.1 |
| Red cell distribution width; RDW (%) | LBXRDW |  |  | 12.9 | 1.3 | 12.6 |
| Platelet count (x10<sup>3</sup> /mm3) | LBXPLTSI |  |  | 240.1 | 64.9 | 232.0 |
| Mean platelet volume; MPV (fL) | LBXMPSI |  |  | 7.9 | 0.9 | 7.9 |
| C reactive protein; CRP (mg/L) | LBXCRP |  |  | 0.4 | 0.8 | 0.2 |
| Total ckolesterol (mg/dL) | LBXTC |  |  | 195.8 | 40.8 | 193.0 |
| HbA1c (%) | LBXGH |  |  | 5.8 | 1.0 | 5.5 |

Footnote: General characteristics of the cases and summary of their laboratory data in the study. \*;
Abbreviation for the variable in the NHANES database, \*\*; Weight status as perceived by the respondent,
\*\*\*; not eligible.

### CatBoost Machine Learning Model to Predict Cancer or CHD

The CatBoost machine learning model developed to predict cancer yielded a moderate Area Under the Curve (AUC) value of 0.76, while the model designed to predict CHD performed better, achieving a higher AUC value of 0.87. These AUC values, which reflect the model’s ability to distinguish between positive and negative cases for both cancer and CHD, are visualized in Figure 1. Additionally, the sensitivity figure, also known as recall, that measures the proportion of actual positive cases that are correctly identified by the model, indicating its ability to detect true positives, was calculated to be 0.67 for cancer and 0.78 for CHD, by using 13 and 12 predictive features respectively, as selected by the RFECV algorithm. A summary of efficacy metrics, revealed by the CatBoost model, for both cancer and CHD predictions is provided in Table 2.

**Figure 1:**
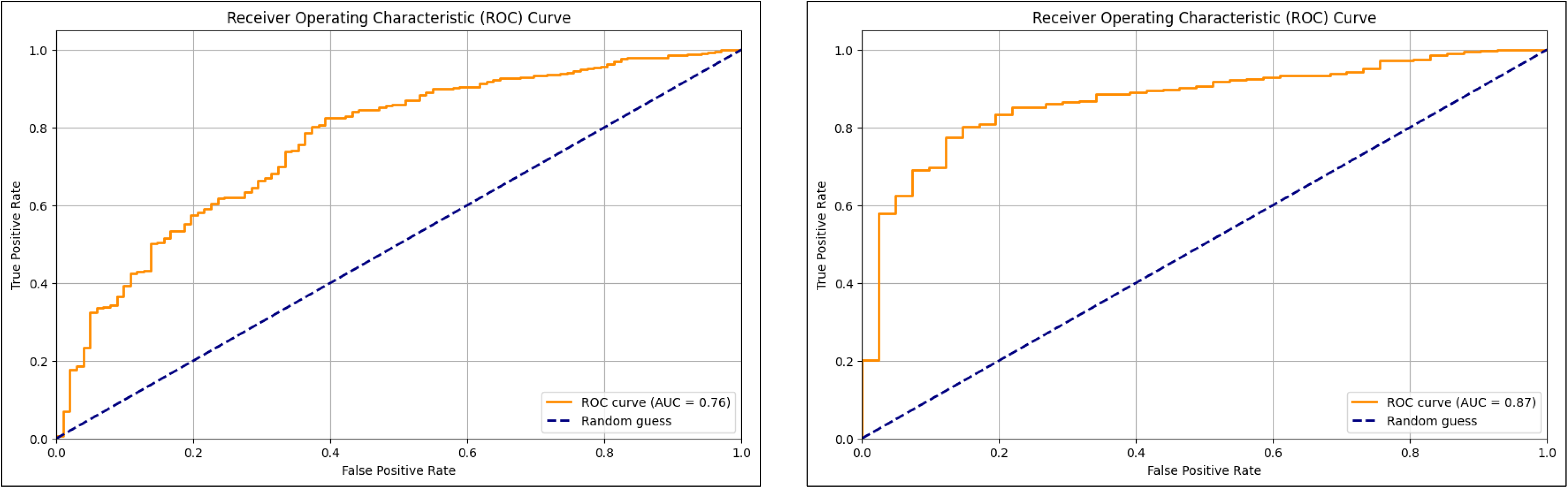
ROC Curves for cancer and coronary heart disease (CHD) occurrence. 1a. ROC curve for binary classification of cancer. 1b. ROC Curve for binary classification of CHD. The curve plots the true positive rate (sensitivity) against the false positive rate (1 - specificity) at various threshold settings. An AUC value closer to 1 representing a better classification model. The dashed diagonal line represents the performance of a random classifier (AUC = 0.5).

**Table 2.** Performance metrics of the cancer and CHD prediction models.

| Endpoint | AUC | Accuracy | Sensitivity | Specificity | Precision | F1 score |
| --- | --- | --- | --- | --- | --- | --- |
| Cancer | 0.76 | 0.70 | 0.67 | 0.70 | 0.20 | 0.31 |
| Coronary heart disease | 0.87 | 0.83 | 0.78 | 0.83 | 0.14 | 0.22 |

Footnote: AUC; area under curve, $\text{Accuracy} = \frac{TP+TN}{TP+TN+FP+FN}$ , where True Positives (TP): correctly predicted positive cases, True Negatives (TN): correctly predicted negative cases, False Positives (FP): incorrectly predicted positive cases (actual negatives predicted as positives), False Negatives (FN): incorrectly predicted negative cases (actual positives predicted as negatives). $\text{Sensitivity} = \frac{TP}{TP+FN}$ , $\text{Specificity} = \frac{TN}{TN+FP}$ , $\text{Precision} = \frac{TP}{TP+FP}$ , $\text{F1 Score} = 2 \times (\text{Precision} \times \text{Sensitivity}) / (\text{Precision} + \text{Sensitivity})$ .

### Feature Importance of Predictor Variables in the CatBoost Model

The optimal number of predictor variables that were selected by the RFECV algorithm and maximized the performance of the CatBoost models, in terms of sensitivity, specificity, and accuracy, were identified as 13 for cancer and 12 for CHD. In the cancer model, the 5 most influential features were age, gender, financial status (income from interest, dividends, or rentals), Neutrophil to Lymphocyte Ratio, and HbA1C levels., with mean SHAP values of 0.84, 0.15, 0.11, 0.08 and 0.04, respectively. In the CHD model, the top 5 predictors were age, gender, platelet count, family history of CHD, and Red Cell Distribution Width, with mean SHAP values of 1.60, 0.46, 0.32, 0.28, and 0.21, respectively. Refer to Figure 2 for mean absolute SHAP values for the cancer and CHD outcomes.

**Figure 2:**
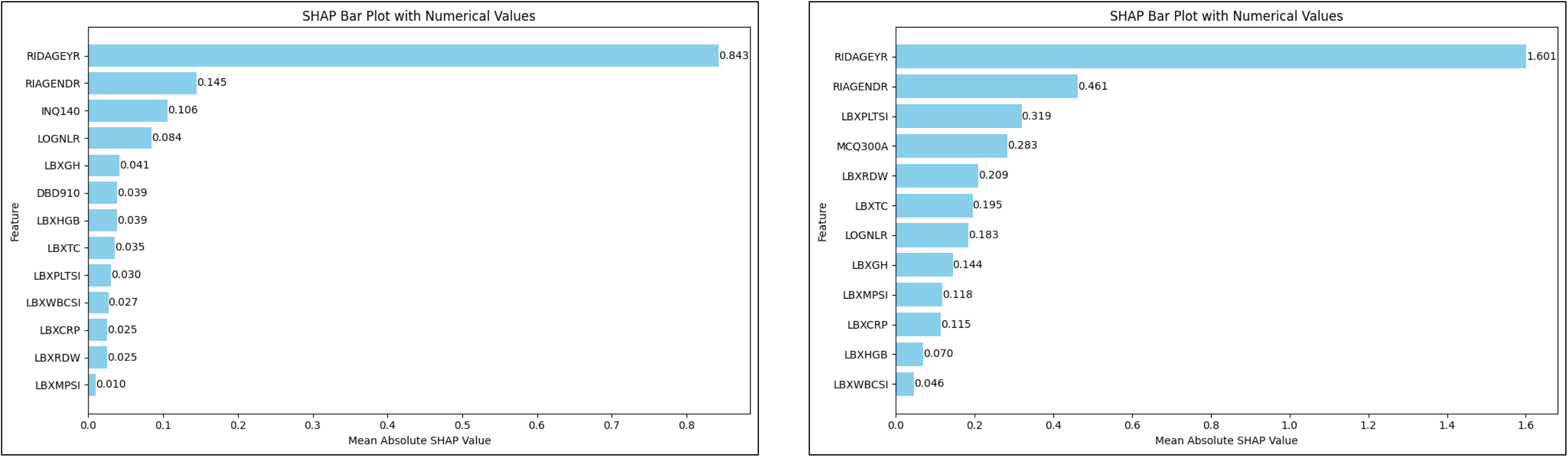
Mean absolute SHAP value plots for cancer and coronary heart disease classification using the CatBoost model. 2a. Mean SHAP value plot for cancer classification. 2b. Mean SHAP value plot for CHD classification. The x-axis represents the mean absolute SHAP values for each feature, showing their contribution to the model’s prediction of cancer, or CHD. The plots illustrate the key predictors in the model. RIDAGEYR; age, RIAGENDR; gender, DBD910; diet habits, INQ140; income, MCQ300A; relative with heart attack, LBXWBCSI; white blood cell count, LBXHGB; hemoglobin level, LBXRDW; red cell distribution width, LBXPLTSI; platelet count, LBXMPSI; mean platelet volume, LBXCRP; C reactive protein, LBXTC; total cholesterol, LBXGH; HbA1c, NLR; neutrophil to lymphocyte ratio. Also, refer to Table 1 for more information about the predictors used in this study.

Among the top 5 predictors for cancer; older age (in cases with and without cancer: 65 versus 48), female gender (52% versus 50%), higher income (44% versus 26%), higher neutrophil to lymphocyte ratio (logarithmic transformation; 0.35 versus 0.30), and higher glycosylated hemoglobin (HbA1c; 5.86 versus 5.74) levels were associated with an increased likelihood of developing cancer. In addition, for CHD, older age (in cases with and without CHD: 69 versus 49), male gender (73% versus 27%), %), lower platelet counts (208000 versus 242000/mm3), a family history of heart attack (27% versus 12), and higher red cell distribution width (RDW; 13.5% versus 12.9%) levels were identified as key risk factors, constituting the 5 top predictors, for the occurrence of CHD. Figure 3 displays the association directions of the top 3 individual features with cancer and CHD.

**Figure 3:**
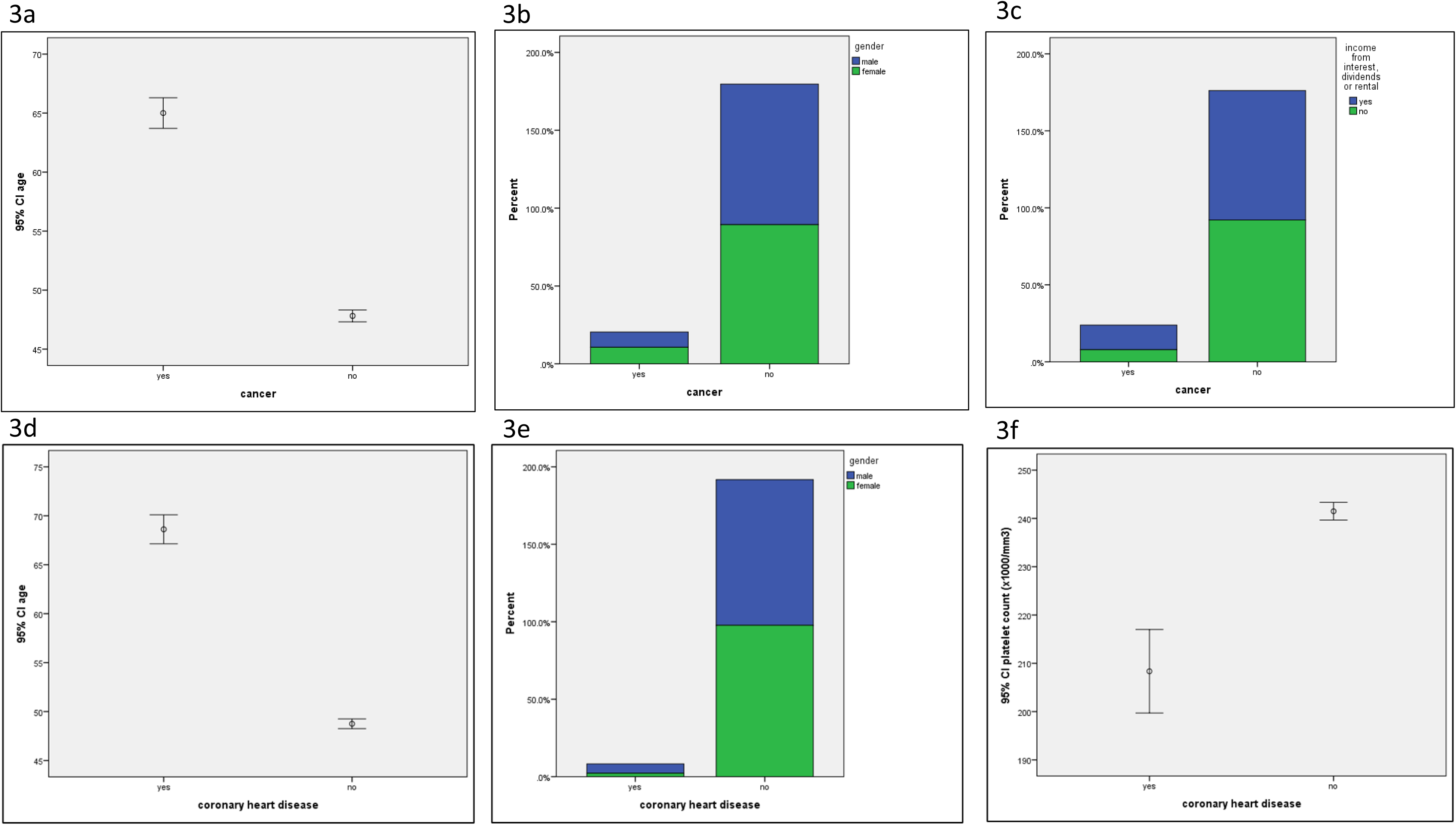
Plot for cancer and CHD related features. 3a. Age for cancer. 3b. Gender for cancer. 3c. Income for cancer. 3d. Age for CHD. 3e. Gender for CHD. 3f. Platelet count for CHD. Top 3 predictor features for cancer and CHD occurrence are presented.

## DISCUSSION

This study demonstrated that the primary predictors for cancer and CHD from the NHANES database, as identified through the machine learning models, are largely related to aging, sociodemographic factors, and laboratory measurements. These findings suggest that these variables can be considered predisposing factors or correlates for the development of both cancer and CHD. As a result, the panel of predictors identified in this study has the potential to be used as an initial screening tool for assessing the risk of developing cancer, CHD, or both conditions. However, while our machine learning models achieved reasonable accuracy in predicting both diseases, the precision rates were low, at 20% for cancer and 14% for CHD. This indicates that any positive prediction should be interpreted with caution, as there is a significant chance it could be a false positive. Thus, in theory, while the model may be useful for initial risk assessment, its predictions—particularly positive ones—should be validated through clinical assessments or additional diagnostic testing.

It is well known that approximately 40-50% of the risk for coronary heart disease (CHD) and 10-20% of the risk for common cancers can be attributed to genetic factors [24,25]. Given this strong genetic component, it is logical to consider genetic approaches for predicting cancer or CHD risk in otherwise healthy individuals. Employing such genetic risk models can encourage proactive lifestyle changes—such as improved diet and exercise habits—and enable early prophylactic interventions before disease development. Thus, any predictive model for the prediction of cancer or CHD risk, including ours, is expected to improve in accuracy by the integration of genomic data as additional predictive features.

Genome-wide association studies (GWAS) have identified around 450 high-risk genetic variants linked to various cancers and approximately 160 genome-wide significant loci (P < 5 × 10⁻⁸) associated with CHD [25,26]. These genetic markers offer promising opportunities for identifying individuals at high risk early on, especially as advances in technology make genetic testing more accessible and affordable.

Incorporating genetic data into predictive models could significantly enhance their accuracy. So, as noted above, in the context of our study, integrating genetic information into our machine learning models could improve their predictive power for both cancer and CHD.

In addition to genetic data, non-genetic clinical data have proven effective in quantifying the risk of cancer and CHD. For example, a Support Vector Machine (SVM) model has been used to assess CHD risk, achieving an AUC of 0.89, which demonstrates strong predictive capability using clinical, non-genetic inputs [27]. Similarly, a Gradient Boosting algorithm, utilizing laboratory, demographic, and comorbidity data, reported an AUC of 0.761, further validating the utility of clinical data in predicting CHD risk [28]. In the field of lung cancer detection, machine learning models have also shown substantial promise. One machine learning algorithm applied in the National Lung Screening Trial demonstrated an AUC of 0.797 in the validation set, with a sensitivity of 0.830, indicating its effectiveness in detecting lung cancer cases [29]. These examples underscore the potential of non-genetic, data-driven approaches, similar to mine, for identifying individuals at risk of these diseases.

Aging is known to be associated with chronic inflammation, mediated by cytokines, which is one of the causal factors in the development of atherosclerosis and cancer. Chronic inflammation is a shared factor contributing to both cancer and CHD [30]. Other related factors, such as lymphopenia, leukocytosis, and elevated glycosylated hemoglobin levels, may also occur alongside chronic inflammation and metabolic syndrome. These factors emerged as important predictors in our study, reinforcing their role in the pathogenesis of both cancer and CHD [31,32]. These findings highlight the interconnectedness of inflammation, metabolic syndrome, and hematologic changes in the development of these conditions, justifying their use as predictive markers in machine learning models.

In our study, the CatBoost machine learning algorithm exhibited performance metrics that were comparable to other models reported in the literature. The AUC values for cancer and CHD were 0.76 and 0.87, respectively, demonstrating a reasonable level of accuracy. These results are consistent with AUC values reported in prior studies (28, 29). However, despite the strong performance in terms of AUC, our models yielded a substantial proportion of false positives in the predictions, which limits the clinical utility of the models. Nevertheless, our models successfully ranked and compared the importance of various features, and this may prove useful in future longitudinal studies. With the inclusion of additional cases and features, more accurate and discriminative predictive models could be developed for cancer and CHD. This study adds to the growing body of literature in medicine, where machine learning models can be useful in early prediction.

One significant limitation of our study is the nature of the NHANES database and the way participants are recruited. NHANES is not a longitudinal study, meaning it does not involve long-term follow-up of participants. Each survey cycle is cross-sectional, capturing data at a single point in time from a new representative sample for that cycle. As a result, cancer or CHD diagnoses in the dataset represent the presence of these conditions at the time of data collection, rather than reflecting a prospective observation of disease development over time. This makes it challenging to interpret our results fully for several reasons. First, causal associations cannot be inferred because the data lack a temporal dimension. Second, some of the predictor variables may be influenced by the outcomes (i.e., the presence of cancer or CHD) rather than serving solely as risk factors.

The primary objectives of this study were twofold: to compare the feature importance of predictor variables and to test the utility of a general machine learning algorithm in predicting the occurrence of cancer and CHD. From the perspective of feature importance, I successfully ranked the relevant predictors from the NHANES database for cancer and CHD occurrence. Although I developed reasonably performant binary classification models for both conditions, I acknowledge that their clinical utility is limited by the low Precision Rate and the non-temporal nature of the data. Future studies incorporating longitudinal and genetic data, involving many thousands of cases, could be analyzed using machine learning or neural network models to develop more accurate algorithms for the prediction of cancer and CHD. The development of such models would be promising from a public health perspective.

## Data Availability

All data produced in the present study are available upon reasonable request to the authors

https://www.cdc.gov/nchs/nhanes/index.htm

## KEY MESSAGE

Various personal, demographic, and laboratory features can predict the occurrence of cancer or CHD. Applying machine learning to larger datasets—especially those incorporating genomic features—could lead to more accurate algorithms for classifying individuals at high risk of developing these diseases. Machine learning algorithms, using a methodology similar to that of this study, could be highly valuable for screening purposes.

## DECLERATIONS

### Ethics approval and consent to participate

None required as only online open dataset has been used. Consent for publication: None required as only online open dataset has been used.

### Availability of data and materials

Available upon reasonable request. The open database for the NHANES study was used to collect data through its official website with the following web address; “https://www.cdc.gov/nchs/nhanes/index.htm”.

### Funding

No funding has been received for this study.

### Authors’ contributions

Hakan Şat Bozcuk found the research question, wrote the Python code, collected data, did the statistical analysis, wrote the draft and edited before submission.

## Acknowledgements

None.

## REFERENCES

1. CDC. Heart disease risk factors. Available from: https://www.cdc.gov/heart-disease/risk-factors/index.html. Accessed 27/10/2024.

2. World Health Organization. Cancer. Available from: https://www.who.int/news-room/fact-sheets/detail/cancer. Accessed 27/10/2024.

3. Koene RJ, Prizment AE, Blaes A, Konety SH. Shared Risk Factors in Cardiovascular Disease and Cancer. Circulation, 2016; 133(11): 1104–1114.

4. Topol EJ. High-performance medicine: the convergence of human and artificial intelligence. Nature Medicine, 2019; 25(1): 44–56.

5. Bindra S, Jain R. Artificial intelligence in medical science: a review. Irish Journal of Medical Science, 2024; 193:1419–1429.

6. Yala A, Lehman C, Schuster T, Portnoi T, Barzilay R. A deep learning mammography-based model for improved breast cancer risk prediction. Radiology, 2019; 292(1): 60–66.

7. Bozcuk HŞ. Machine learning assisted web application for identifying beneficial drug candidates for genetic alterations in cancer patients. MedRxiv. Available from: https://www.medrxiv.org/content/10.1101/2024.06.03.24308392v1. Accessed 27/10/2024.

8. Bozcuk HŞ, Şen AE, Artaç M, Kaya B. Unveiling brain region patterns in PET/CT scans for lung cancer assessment: a computational AI framework. Journal of Cancer Science and Clinical Therapeutics, 2023; 7: 179–185.

9. Wang HL, Hsu WY, Lee MH, et al. Automatic machine-learning-based outcome prediction in patients with primary intracerebral hemorrhage. Frontiers in Neurology, 2019; 10. 10.3389/fneur.2019.00910.

10. Hancock JT, Khoshgoftaar TM. CatBoost for big data: an interdisciplinary review. J Big Data, 2020; 7:94. 10.1186/s40537-020-00369-8.

11. CDC. National Health and Nutrition Examination Survey. Available from: https://www.cdc.gov/nchs/nhanes/index.htm. Accessed 27/10/2024.

12. Google Colaboratory. Available from: https://colab.google/. Accessed 27/10/2024.

13. Pandas. Available from: https://pandas.pydata.org/. Accessed 27/10/2024.

14. Gazoni E, Clark J. Openpyxl - A Python library to read/write Excel 2010 xlsx/xlsm files. Available from: https://openpyxl.readthedocs.io/en/stable/. Accessed 27/10/2024.

15. IBM SPSS Statistics. Available from: https://www.ibm.com/products/spss-statistics. Accessed 27/10/2024.

16. RFECV. Available from https://scikit-learn.org/stable/modules/generated/sklearn.feature_selection.RFECV.html. Accessed 27/10/2024.

17. A guide to F1 score. Logunova I. Available from https://serokell.io/blog/a-guide-to-f1-score. Accessed at 27/10/2024.

18. Welcome to the SHAP documentation. Available from: https://shap.readthedocs.io/en/latest/. Accessed at 27/10/2024.

19. Nahm FS. Receiver operating characteristic curve: overview and practical use for clinicians. Korean J Anesthesiol 2022; 75(1): 25–36.

20. Numpy. Available from: https://numpy.org/. Accessed at 27/10/2024.

21. Scikit-learn. Machine learning in Python. Available from: https://scikit-learn.org/stable/. Accessed 27/10/2024.

22. CatBoost is a high-performance open-source library for gradient boosting on decision trees. Available from: https://catboost.ai/. Accessed at 27/10/2024.

23. Matplotlib: Visualization with Python. Available from: https://matplotlib.org/. Accessed at 27/10/2024.

24. Won H-H, Natarajan P, Dobbyn A, Jordan DM, Roussos P, Lage K, Raychaudhuri S, Stahl E, Do R Disproportionate contributions of select genomic compartments and cell types to genetic risk for coronary artery disease. PLoS Genet 2015;11(10):e1005622.

25. Sud A, Kinnersley B, Houlston RS. Genome-wide association studies of cancer: current insights and future perspectives. Nature Reviews Cancer, 2027; 17: 692–704.

26. Van der Harst P, Verweij N. Identification of 64 novel genetic loci provides an expanded view on the genetic architecture of coronary artery disease. Circ Res, 2018; 122(3): 433–443.

27. Haq AU, Li JP, Memon MH, et al. A hybrid intelligent system framework for the prediction of heart disease using Machine Learning algorithms. Mob Inf Syst, 2018; Article ID 3860146. 10.1155/2018/3860146.

28. Weng SF, Reps J, Kai J, et al. Can machine-learning improve cardiovascular risk prediction using routine clinical data? PLoS ONE 12(4): e0174944.

29. Tammemagi MC, Katki HA, Hocking WG, et al. Selection criteria for lung-cancer screening. N Engl J Med 2013; 368: 728–736.

30. Koelman R, Ramich OP, Pfeiffer AFH, et al. Cytokines for evaluation of chronic inflammatory status in ageing research: reliability and phenotypic characterization. Immunity & Ageing, 2019; 16:11.

31. Yan Y, Gao R, Zhang S, et al. Hemoglobin A1c and angiographic severity with coronary artery disease: A cross-sectional study. Int J Med 2022; 15: 1485–1495.

32. Cyr KJ. What do high Neutrophils and low lymphocytes mean? Verywell Health. Available from: https://www.verywellhealth.com/what-does-high-neutrophils-low-lymphocytes-mean-5210245. Accessed at 27/10/2024.

